# High-Fidelity Measurement of Pulse Arrival Time in Critically Ill Children Using Standard Bedside Monitoring Equipment

**DOI:** 10.1101/2025.03.31.25324979

**Authors:** Ian Ruffolo, Asad Siddiqui, Binh Nguyen, Will Dixon, Azadeh Assadi, Robert Greer, Steven Schwartz, Michael Brudno, Alex Mariakakis, Andrew Goodwin

## Abstract

Pulse arrival time (PAT) is known to be correlated with blood pressure. Although PAT can be measured using electrocardiography (ECG), photoplethysmography (PPG), and other signals commonly available in clinical settings, recent literature has noted that devices recording these waveforms are often subject to many hardware-specific factors related to digital filtering, clock synchronization, temporal resolution, and latency. These factors can introduce relative timing errors between the ECG and PPG signals, resulting in a situation where traditional approaches for PAT measurement will not work as intended. In this work, we propose a methodology that accounts for these confounding factors and generates precise measurements of PAT using standard bedside monitoring equipment. This technique involves using heart rate variability to match heartbeats across waveforms and experimentally profiling the timing systems of bedside medical devices to correct various timing-related artifacts. To improve the precision of the resulting PAT measurements, we model temporal uncertainties stemming from the finite temporal resolution of the waveform samples. We apply this approach to a dataset with roughly 1.6 million hours of continuous ECG and PPG data from over 10,000 unique patients at a pediatric intensive care unit (ICU). After demonstrating that the observed timing artifacts are consistent across the entire dataset, we show that accounting for them results in more reasonable distributions of PAT measurements across age groups. It is our hope that this work will spur discussion around the standardization of PAT measurement using routinely collected signals in a clinical environment.

## 1 Introduction

Blood pressure (BP) is a critical vital sign for monitoring patient health status, reflecting the adequacy of blood flow to vital organs and serving as an essential indicator of cardiovascular function. Abnormalities in BP as a result of various conditions (e.g., septic or hemorrhagic shock) can lead to increased risk of morbidity and mortality due to impacts on vital organ perfusion^1,2^. In high-acuity settings such as the operating room or an intensive care unit (ICU), patients can experience significant physiologic derangement that can lead to rapid changes in BP, making real-time and continuous BP measurement essential for detecting and managing instability. This data is typically obtained using peripherally inserted catheters in the arteries, which is not always feasible or risk-free^3^. Meanwhile, non-invasive blood pressure measurements using a manual or automatic sphygmomanometer provide intermittent information and may not always be accurate. As such, there has been growing interest in the development of noninvasive approaches to continuous BP monitoring^4^.

One such method relies on the measurement of pulse arrival time (PAT) or pulse transit time (PTT), both of which describe the time it takes for the pulse to travel from one measurement site to another within the body. Both PAT and PTT are inversely proportional to pulse wave velocity (PWV), which itself is highly correlated with blood pressure^5–8^. Hospitalized patients are routinely monitored using standard bedside monitoring equipment that collects signals like electrocardiography (ECG) and photoplethysmography (PPG). These signals enable noninvasive measurement of the pulse wave at different sites, such as the heart for ECG and a peripheral site like a fingertip or toe for PPG^9^.

Large datasets of physiological waveforms have become more common in recent years^10–15^, leading many researchers to investigate ways of estimating BP using PAT as a feature in data-driven models^6,16–29^. However, the temporal integrity of the ECG and PPG waveforms in these databases has often been taken for granted. Signals collected from bedside monitoring equipment and clinical data aggregation systems can contain timing errors resulting from clock drift, digital filtering, and signal buffering^30^. For example, Lee et al. observed a delay of approximately 350 ms between the ECG and PPG signals in the VitalDB database^21^. This issue is so salient that documentation for the often-used MIMIC dataset states that it is not designed for inter-waveform analysis^31^, with Liang et al. stating *“one of the next steps is to investigate the synchronicity over the asynchronous MIMIC database”*^32^. Pulse oximetry from different models or manufacturers may preprocess PPG signals in different ways^33^, and medical devices may also alter the signals by applying proprietary pre-processing algorithms^34^, potentially rendering the data unsuitable for some applications^35^.

Ignoring the potential for offsets between the ECG and PPG signals may result in PAT values that are outside a physiologically plausible range. Some researchers have simply filtered out unusual PAT values to use values within what they believe to be a plausible range^25,36^, but this practice has been inconsistently applied across analyses and has likely led to biased results. Therefore, standardized approaches to measurement must be developed to provide a foundation for research and analysis involving PAT.

To the best of our knowledge, nobody has deeply investigated and addressed the multitude of system-related timing errors that affect noninvasive PAT measurement. In this paper, we present a methodology for quantifying and correcting the discrepancies between ECG and PPG signals collected from standard bedside monitoring equipment. We apply these techniques to a large, retrospective dataset of physiological waveforms collected from over 10,000 critically ill children admitted to an ICU. We first utilize this wealth of data to support our assertion that these errors are derived from the system’s hardware. We then compare the distribution of PAT measurements before and after applying our methodology to demonstrate that addressing system-related timing errors is necessary to extract clinically meaningful data.

## 2 Background

### 2.1 Naïve PAT Measurement

PAT measurement requires the identification of the timestamps of fiducial points in both the ECG and the PPG signals. The R-peak is typically used as the timestamp of beat origination according to ECG because of its prominence relative to other components in the waveform, while in PPG, a wide range of possible choices for fiducial point are available^37–39^.

If the ECG and PPG signals are perfectly synchronized, it is reasonable to assume that every beat in the ECG signal can be paired with the following beat in the PPG signal as PAT is being measured. This pairing, denoted as *PAT*_0_ in Figure 1, is the default way of measuring PAT. However, the existence of system-related timing errors between the ECG and PPG signals can break this assumption, meaning that ECG beats should not necessarily be paired with the following beat in the PPG signal. Instead, they should possibly be paired with one further away in time; such possible pairings are denoted *PAT*_1_, …, *PAT*_*n*_ in Figure 1. This issue is particularly salient when the delay in the PPG timestamps is large and/or when the heart rate is high. The remainder of this section outlines several factors that can result in errors in the PPG timestamps.

**Figure 1.**
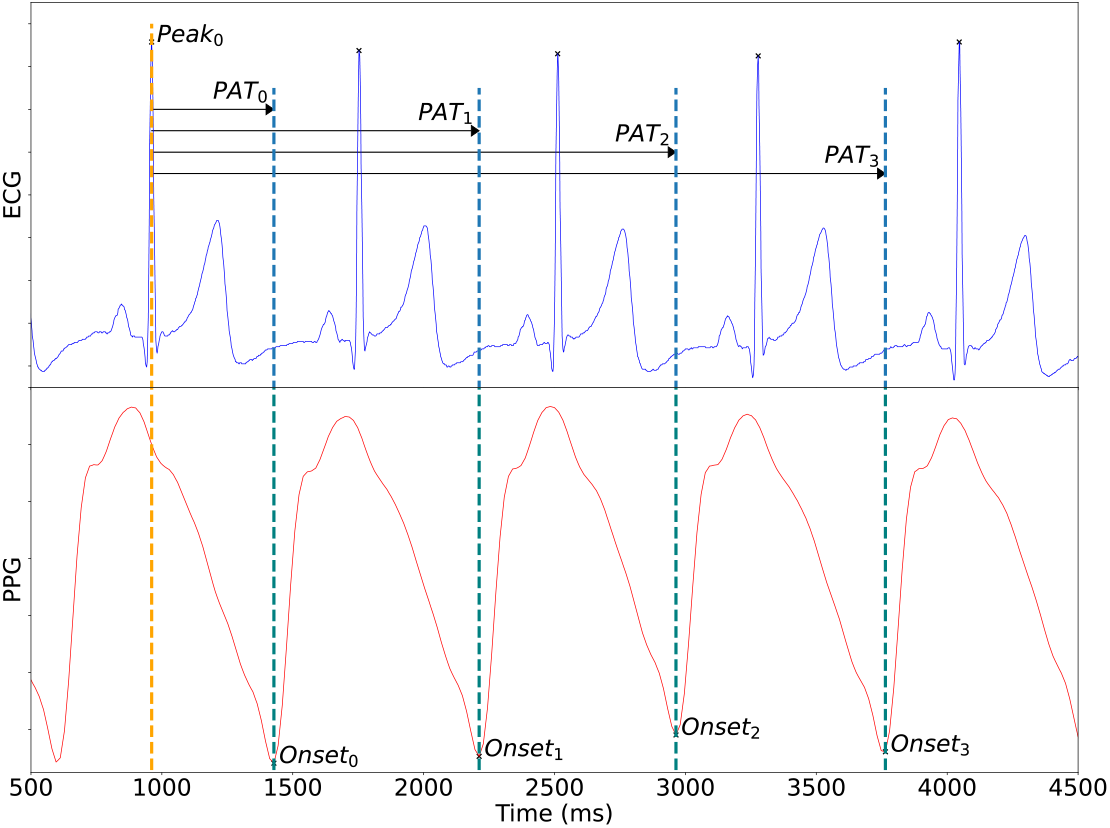
This figure shows the potentially incorrect measurement of PAT from ECG and PPG signals. The R-peak from the first ECG heartbeat (labeled *Peak*_0_) is selected as the first of two fiducial points that are required for the PAT calculation. Four possible PAT values are displayed, *PAT*_0_, *PAT*_1_, *PAT*_2_, and *PAT*_3_ measured to the 1^st^, 2^nd^, 3^rd^, and 4^th^ heartbeat onsets in the PPG respectively. The typical measurement method for calculating PAT seen in prior work naively selects *PAT*_0_ (i.e., the “next beat” in the PPG). However, characterization and correction of the system-related delays in the PPG timestamps will generally shift the PPG signal to the left. This results in a situation where, prior to synchronizing these signals, R-peaks may not be physiologically associated with the “next” beat in the PPG signal.

### 2.2 Sources of PAT Measurement Error

PAT measurement is subject to many sources of error. Some of these errors can be attributed to algorithmic deficiencies like poor fiducial point detection, yet there are others that depend on the software and hardware used to record and collate signals prior to that step. Our dataset comes from an ICU setting where ECG and PPG signals are collected using hardware from two different device manufacturers connected to a bedside monitor (see Section 3.1 for more details). We describe the contributors to the timing errors in this scenario below:

#### 2.2.1 *Sample Rate Imprecision and Re-Synchronization* (Delay_sync_)

After being timestamped by their respective hardware, third-party sensor devices pass their signals through a module connected to the bedside monitor, which then reassigns timestamps according to the monitor’s clock. Since the sample rates supplied by both device manufacturers are not precisely specified, there is a non-zero relative drift rate between the timestamps that are generated when using the nominal sample frequencies^40,41^. This relative drift rate is periodically corrected by synchronizing the two signals, but the continuous drift-and-correct cycle leads to a sawtooth-shaped series of timing errors observed by other researchers^42–44^; an example of this artifact is shown later in Panel B of Figure 2. Upon observing this artifact, Bennis et al. suggest, *“This sawtooth, together with a large module-dependent absolute difference in PTT, renders the thus-derived PTT insufficient for clinical purposes”*^*44*^. If the signal is passed through multiple devices (e.g., an extension module before going to the monitor), multiple sawtooth artifacts could be present within the same signal.

**Figure 2.**
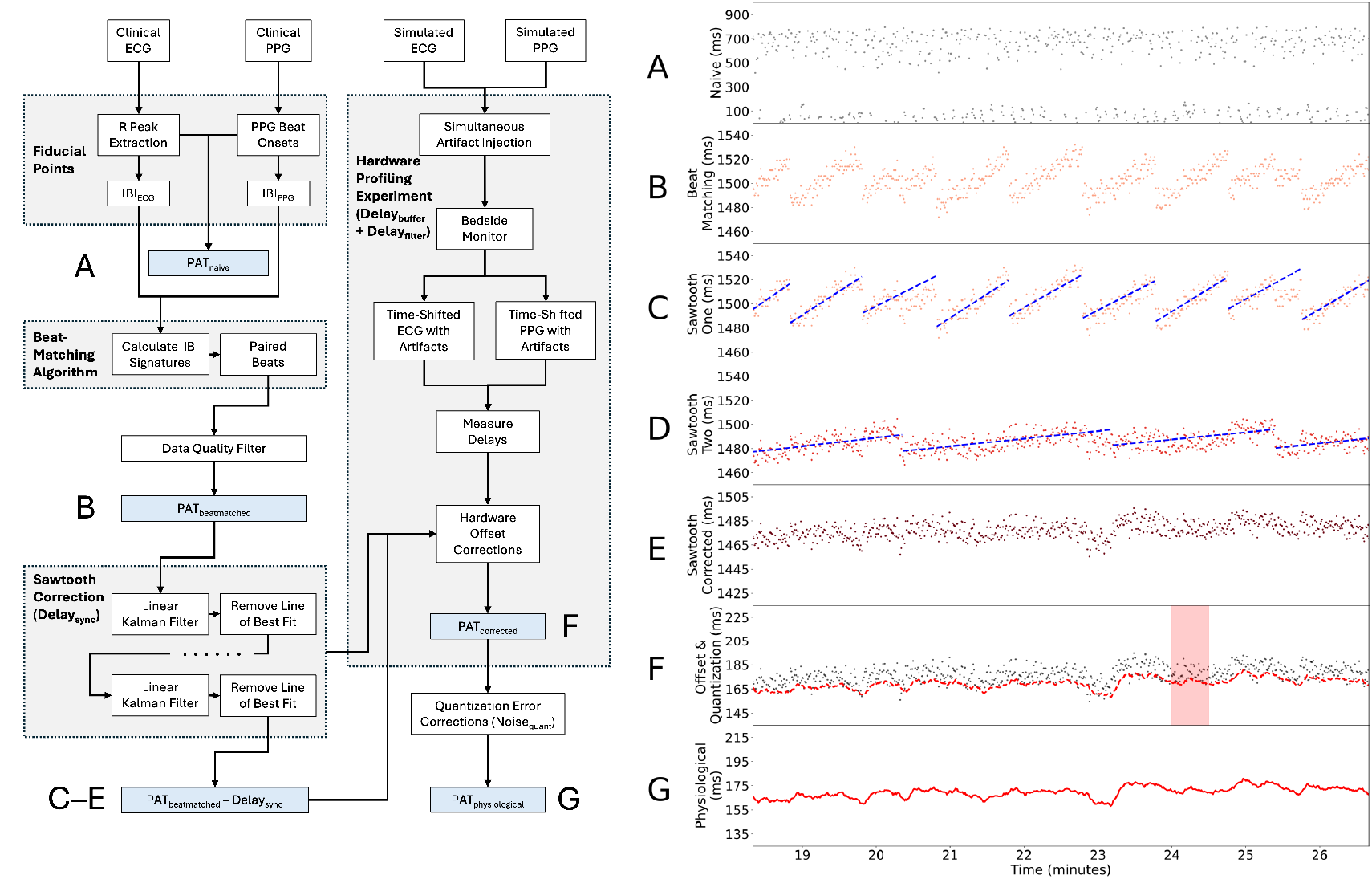
The left side of the figure provides an overview of the methodology we employed for precise PAT measurement, while the right side shows the results of this procedure on a real-world PAT timeseries. (A) Naïve PAT measurements are taken between the ECG and the PPG assuming that ECG R-peaks should be matched with the “next beat” in the PPG. (B) *PAT*_*beatmatched*_ is calculated using our beat-matching algorithm. (C) A Kalman filter is used to identify the sawtooth artifact that appears due to clock drift and synchronization. (D) After removing the first sawtooth artifact, a second one appears in our dataset due to multi-stage hardware. (E) *PAT*_*beatmatched*_ − *PAT*_*sync*_ accounts for the cumulative effects of the sawtooth artifacts. (F) *PAT*_*corrected*_ excludes *Delay*_*bu f f er*_ and *Delay*_*filter*_, which are both derived from hardware profiling. (G) *PAT*_*physiological*_ is the latent PAT signal that addresses measurement errors resulting from the finite temporal resolution of the fiducial points.

**Figure 3.**
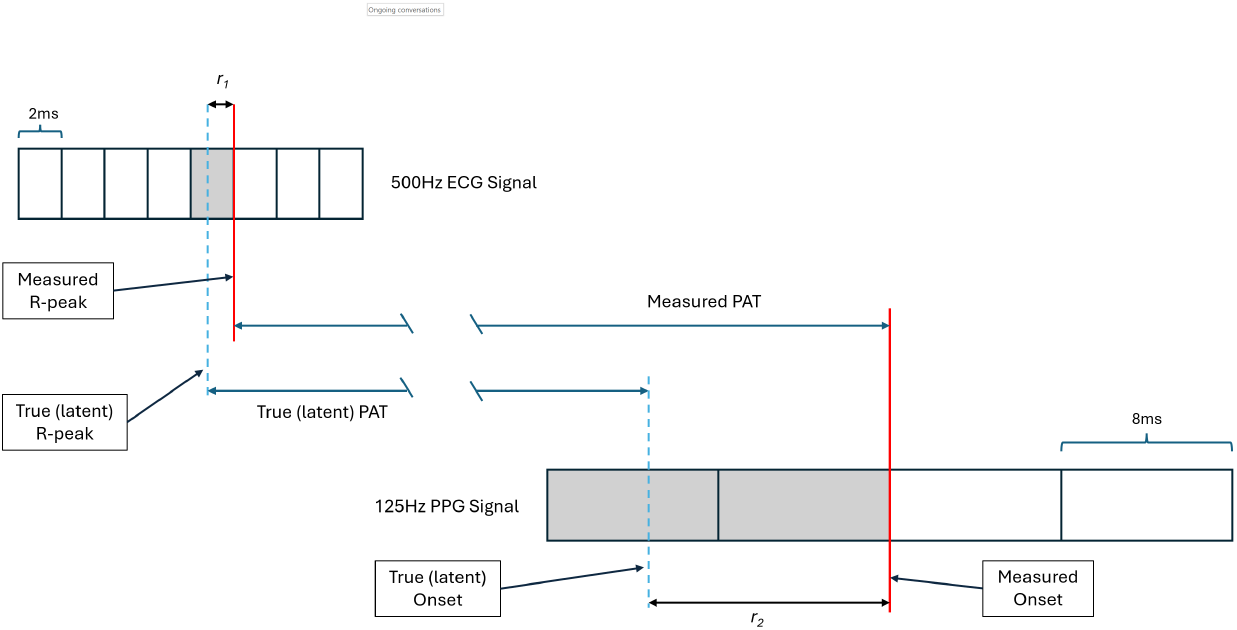
The impact of quantization on the discrepancy between measured PAT and *PAT*_*physiological*_. While the temporal resolution of fiducial points in the 500-Hz ECG is 2 ms, the effective resolution of the 125-Hz PPG is 16 ms when PAT is being measured.

#### 2.2.2 Filter-Based Delays Delay_filter_

Clinical monitors typically apply digital filters to signals in an attempt to enhance signal quality for downstream visualization and analysis^45,46^. PPG signals particularly require aggressive averaging and smoothing due to the amount of noise in the raw signals^47^. A byproduct of this process is delays and other timing errors that can impact the relative timing of fiducial points within the PPG signal itself and with respect to other signals^48–51^. The characteristics of these filters are often considered proprietary and are likely to vary across different medical devices. This means that delays of different magnitudes may be present in signals collected from different medical device manufacturers.

#### 2.2.3 *Digital Buffering* (Delay_buffer_)

The main focus of a monitoring device is to provide a steady output visualization to facilitate clinical care. To this end, many sensors, clinical monitors, and data aggregation systems may buffer measurements to avoid pauses and discontinuities in their continuous output^52^. The magnitude of the delays introduced by this buffering process may vary across devices and can be further compounded by the presence of intermediary data aggregation systems^33^.

#### 2.2.4 *Temporal Resolution* (Noise_quant_)

Digital signal quantization and filtering algorithms can limit the temporal resolution of fiducial points that are used to measure PAT. The ECG and PPG signals in our setting are nominally quantized to 2 ms and 8 ms given their respective sampling rates of 500 Hz and 125 Hz. However, we will later explain that the fiducial points in the PPG signals in our dataset actually have a resolution of 16 ms.

### 2.3 Summary

Timestamps in PPG signals can be altered by a range of different effects including clock drift, digital filters, and signal buffering. We will later demonstrate that the magnitude of system-related delays in the PPG signal can exceed 1300 ms. In pediatric populations, interbeat intervals (IBIs) can range from 350 ms to 750 ms^53,54^, while PAT may vary from 100 to 370 ms depending on the size and age of the patient, sensor locations, and other factors^55^. The delays introduced into the PPG signal’s timestamps result in a situation where heartbeats in the ECG and PPG signals can be out of phase by one or more beats, particularly in children with higher heart rates.

In order to correctly measure PAT, we require a method that is capable of matching an R-peak in the ECG with its *corresponding* fiducial point in the PPG signal. Once this match has been found, we can measure the time between the R-peak and the fiducial point in the corresponding beat in the PPG, which we call *PAT*_*beatmatched*_. The beat-matched PAT also includes contributions from the data collection system, which we have categorized as follows:

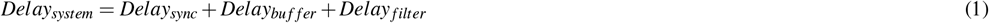

Once these systematic delays have been quantified, they can be subtracted from *PAT*_*beatmatched*_ to leave our corrected PAT as follows:

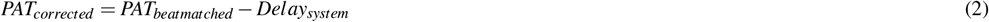

Furthermore, the corrected PAT values are subject to measurement noise as per the following equation:

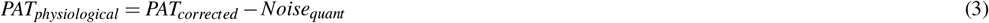

## 3 Methods

The left side of Figure 2 illustrates the various steps in our approach for generating corrected PAT measurements derived from ECG and PPG collected using standard bedside monitoring equipment. The right side of Figure 2 illustrates the results of this process on a real-world PAT timeseries. Once the timing properties of the system have been characterized, this information can be used to correct inter-signal timing errors between the ECG and PPG, thereby synchronizing the two signals. We use these techniques to synchronize signals in our database, thereby allowing us to utilize traditional approaches for PAT measurement. We describe our approach both at a high level and in the context of the specific dataset we analyze in this paper.

### 3.1 Dataset

Physiological waveforms were collected from a 42-bed ICU at the Hospital for Sick Children (SickKids) in Toronto, Canada between 2017 and 2024. This data collection effort was approved by the hospital’s Research Ethics Board (REB#1000068499). The ECG and PPG signals were recorded at nominal frequencies of 500 Hz and 125 Hz respectively. These signals were stored on a secure server in an AtriumDB database^11^. Prior to 2022, signals were collected from Philips IntelliVue MP70 patient monitors coupled with Masimo PPG sensors that were connected through an X2 IntelliBridge device interface. In mid-2022, the monitoring hardware was upgraded to Philips MX750 monitors with an X3 IntelliBridge device interface.

### 3.2 Fiducial Point Detection

We used the findpeaks() function from Neurokit^1^ toolbox to identify R-peaks in the ECG waveform. Meanwhile, we used a method developed by Kavsaouglu et al.^56^ and integrated in the BioSPPy^2^ package to detect the onset of beats in the PPG waveform. While the peak of the pulse wave is also often used as the fiducial point in PPG waveforms, we elected to adopt the onset of each pulse wave since that has been shown to be more reliable^5,37^.

Critically ill children generally have substantially higher heart rates than adults, which required changing the default parameters in these libraries to extend the range of allowable detected heart rates. We adopted an adaptive approach based on the 1 Hz pulse rate signal provided by the Philips patient monitor. If the maximum heart rate from this signal was less than 250 bpm, we used that value as the setting for the max_bpm parameter in BioSPPy, and we used its reciprocal for the mindelay parameter in Neurokit. Otherwise, we set max_bpm to 250 bpm and mindelay to 240 ms. To avoid performing calculations on windows with missed or spurious heartbeats, we removed windows for which the standard deviation in either the ECG or PPG IBIs was greater than 300 ms. We also removed windows for which the difference in mean IBI between the ECG and PPG was greater than 200 ms.

### 3.3 Temporal Quantization of Fiducial Points and IBIs

The temporal resolution of PAT measurements is limited by the sampling rate of the signals^57,58^. There is also a causal relationship between the phenomena being recorded by the signals and the conversion of the digital measurement itself^59^, so we posit that measurements describe events that happened sometime prior to their quantized timestamps. As illustrated in Figure 3, each sample in the 500-Hz ECG signal summarizes the heart’s electrical activity over the preceding 2 ms. This means that any measured R-peaks may have actually occurred up to 2 ms prior. Similar logic would lead one to believe that each sample in the 125-Hz PPG summarizes blood volume changes over the preceding 8 ms. However, a close examination of the PPG IBIs contradicted this expectation.

Figure 4 shows the distribution of differences between consecutive IBIs in the ECG and PPG signals. While the separation between ECG IBIs was always a multiple of 2 ms, the separation between PPG IBIs was highly likely (>95%) to be an even number of samples. In other words, the resolution of the beat onsets extracted from the PPG signal was 16 ms, or two consecutive 8 ms samples. This observation persisted when different techniques were used for fiducial point detection, confirming that this was a property of the signal and not a limitation of algorithms. Therefore, the effective resolution of the pulse onsets in the PPG in our dataset is actually 16 ms when we are measuring PAT.

**Figure 4.**
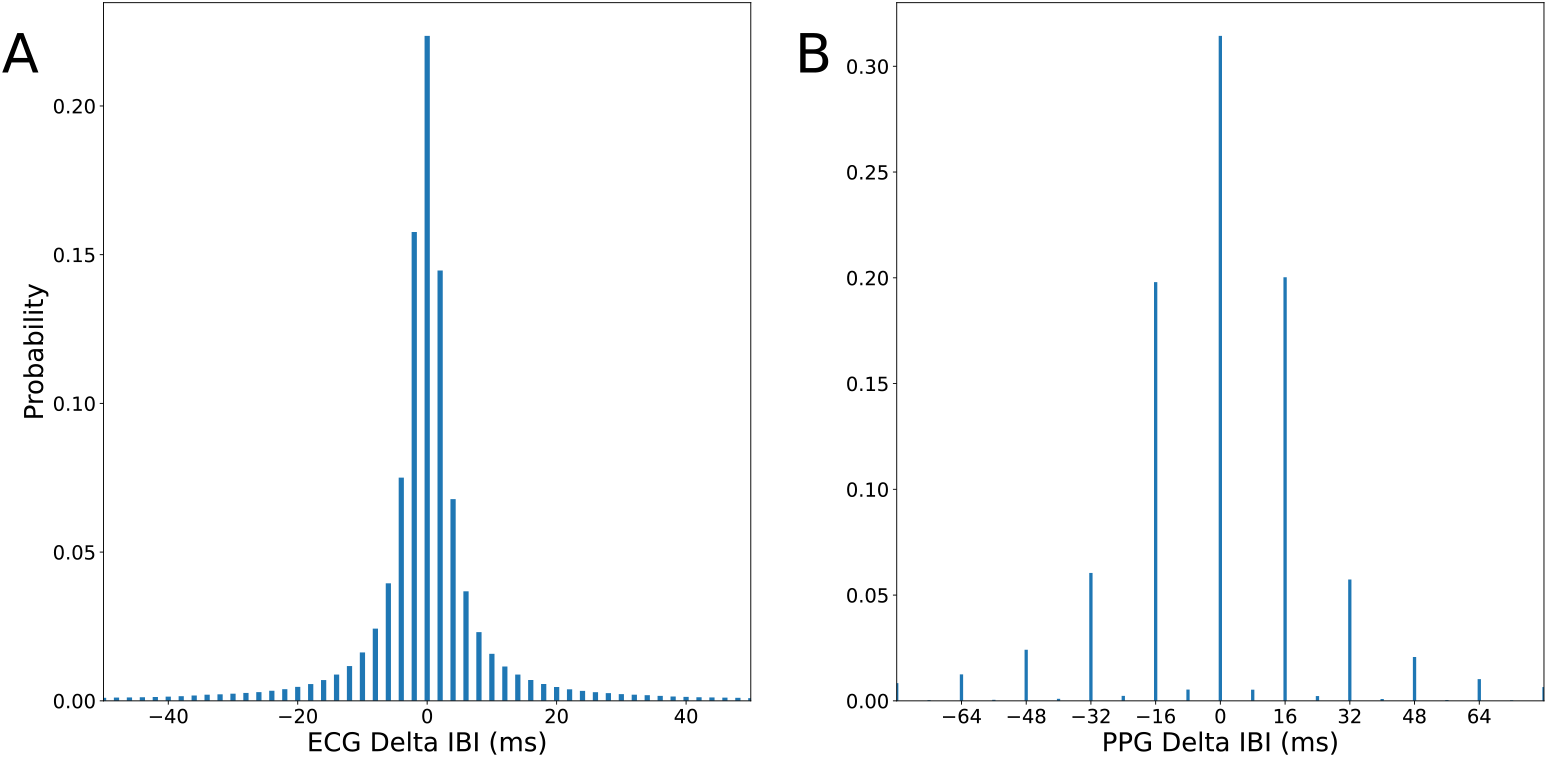
The distribution of differences between consecutive IBIs derived from (A) ECG and (B) PPG.

### 3.4 Beat Matching

We extend the algorithm proposed by Goodwin et al.^60^ to match beats across the ECG and PPG signals. This technique relies on identifying segments with similar sequences of IBIs between the two signals. For a given beat *i*, we generated a unique signature *Sig*_*i*_ = {*IBI*_1_, *IBI*_2_, …, *IBI*_20_} corresponding to the IBIs of the 20 beats after it. We discarded any signatures containing an IBI that would have resulted in an instantaneous heart rate measurement below 50 bpm or above 250 bpm since they were likely to be compromised by missed or spurious heartbeats.

Signatures with more drastic changes in IBI were assumed to be more unique, so we used the following as a score of the signature’s uniqueness:

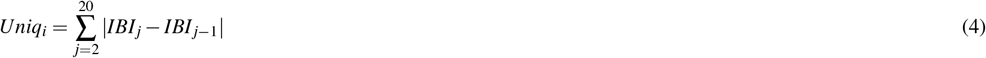

Beyond calculating this score, we discarded signatures for which the 75^th^-percentile of IBI differences falls below the corresponding signals’ quantization (2 ms for ECG, 16 ms for PPG). Signatures that failed this criterion were not necessarily low-quality, but they were challenging to match across signals due to a lack of uniqueness.

We compared the remaining signatures across the two signals according to their Euclidean distance. In our specific implementation, we bounded this search procedure to only compare signatures within 6 beats of one another given empirical observations that inter-signal timing delays typically did not exceed that duration. Matches between signatures with low uniqueness scores were less reliable, so we prioritized matches between vectors with higher IBI variability. Therefore, applying our beat-matching algorithm for a given beat *i* in the ECG entailed minimizing the following expression to identify the

#### Algorithm 1

An abridged version of the linear Kalman filter (KF) technique used to track a sawtooth artifact in a PAT time series

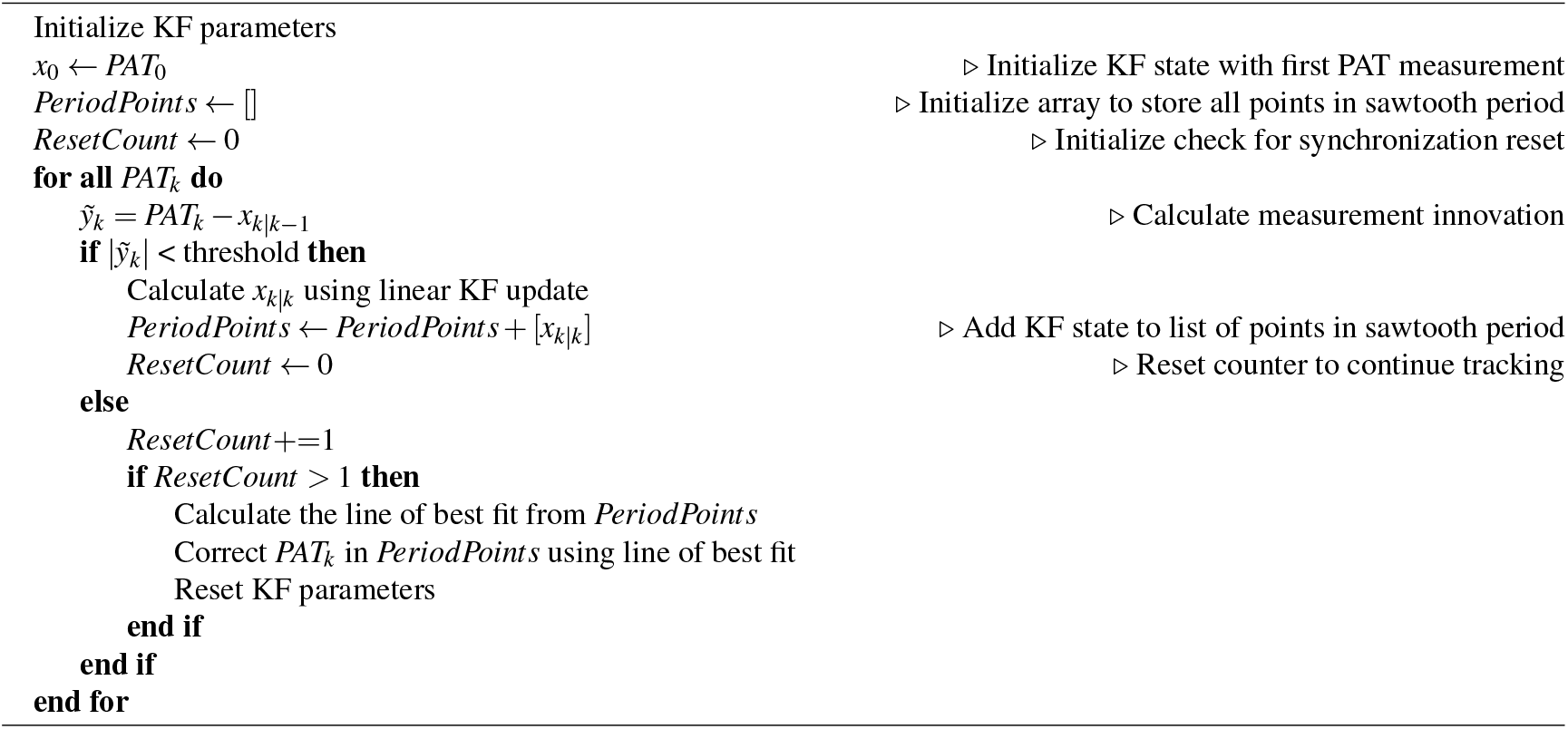

corresponding beat *j* in the PPG:

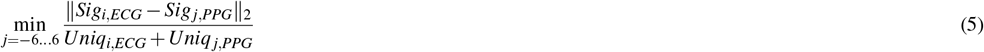

Once all the beats were matched, we calculated the PAT for all the matched beats within a 60-minute frame. The distribution of these beat-matched PATs contained multiple peaks roughly spaced according to the average IBI. We assumed that the most prominent peak corresponds to correctly matched beats, while the remaining peaks contained mismatches. The mismatches could hypothetically be corrected instead of discarded, but for this work, we elected to discard these cases in order to provide the most robust signal for subsequent steps in the analysis. We algorithmically performed this final filtering step by discarding PATs beyond two standard deviations from the frame’s mean.

### 3.5 Characterization of Clock Drift Artifacts

As mentioned in Section 2.2.1, clock drift and re-synchronization can produce artifacts in the PAT time series; in fact, there can be multiple sawtooth artifacts superimposed on one another when signals are passed through multiple interfaces. The periods and amplitudes of these artifacts can be identified via manual inspection of a *PAT*_*beatmatched*_ timeseries. While we used manual inspection to confirm the presence of two sawtooth artifacts in our dataset, we propose an automated approach that estimates the characteristics of (amplitude, period, and phase) of a sawtooth artifact in the presence of physiological variation and other measurement noise.

As outlined in Algorithm 1, the technique centers around a Kalman filter that tracks the linear rise of beat-matched PAT measurements over time. When the difference between the predicted state and a new measurement (i.e., the measurement innovation) exceeds 16-ms, the Kalman filter resets under the assumption that a clock synchronization event has occurred. This threshold is based on the resolution of PATs in our dataset. To make this algorithm robust to outliers, the Kalman filter only resets after two samples exceed the threshold. Once a cycle of the sawtooth artifact has been identified, the estimated measurements are used to fit a line. The line is subtracted from the PAT measurements that were fed into the Kalman filter to suppress the drift during that cycle.

### 3.6 Hardware Profiling

The total *Delay*_*bu f f er*_ +*Delay*_*filter*_ can be inferred if *Delay*_*sync*_ and the expected distribution of PATs are known. Since we did not have access to the latter, we conducted explicit experiments with the hardware at our institution in order to empirically estimate *Delay*_*bu f f er*_ and *Delay*_*filter*_. We first synthetically generated ECG and PPG waveforms representative of cardiac function. This entailed using a PS420 multi-parameter simulator for synthetic ECG and a Masimo SET Tester for synthetic PPG. We then injected simultaneous voltage step changes into both signals before they arrived at the transducers. The voltage changes were introduced every 20 seconds, alternating between stepped increases and decreases in voltage, and this was done out of phase with the heartbeats to avoid coincidental alignment that would affect our observations.

As shown in Figure 5, the voltage step changes appeared at different times in the two signals collected by the monitor. We visually annotated and measured two intervals associated with each artifact. The first was the relative difference between the time that the artifact arrived in each signal (Δ*t*_*arrival*_), while the second was the relative time difference between the introduction of the artifact into the ECG signal and the moment the PPG waveform morphology began to deviate from its baseline pattern (Δ*t*_*deviation*_). These measurements incorporate different combinations of system-related delays:

**Figure 5.**
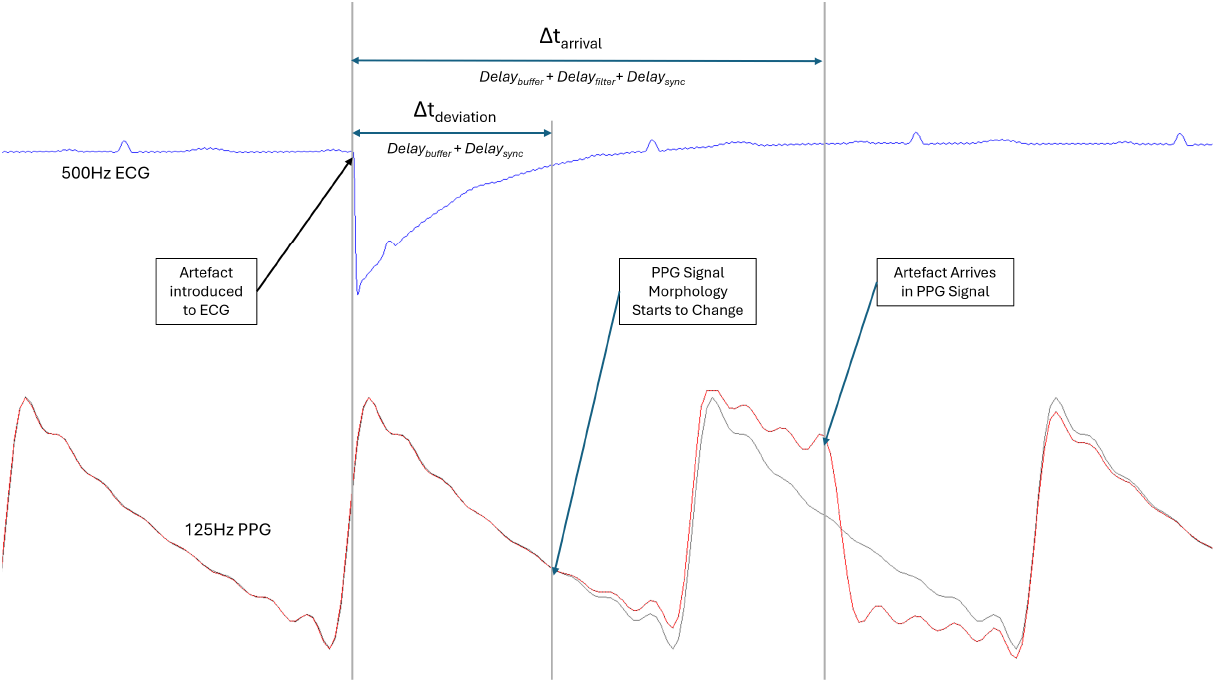
Example output from the hardware profiling experiment. Artificially generated artifacts were simultaneously introduced into the ECG and PPG signals. Δ*t*_*arrival*_ represents the relative time difference between the expression of the artifact in the ECG and PPG signals. Δ*t*_*deviation*_ describes the moment the PPG signal begins to deviate from the repeating pattern in the synthetically generated signal. The ECG and PPG signals were generated using different heart rates, but this did not affect the experiment since the artifact detection process is independent of physiology.

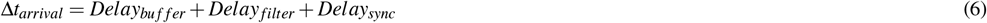

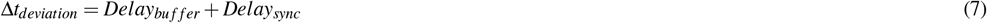

Having previously estimated the properties of *Delay*_*sync*_ using Kalman filtering, we were able to use these relationships to de-convolve the magnitudes of *Delay*_*bu f f er*_ and *Delay*_*filter*_.

### 3.7 Quantization Noise Correction

PAT measurements are subject to the finite temporal resolution of the fiducial points in each signal. To account for these measurement errors, we used a Monte Carlo simulation to model the random processes associated with the quantization of the timestamps (*Noise*_*quant*_). Since the measurements from our hardware profiling experiment are subject to measurement errors stemming from temporal quantization, we executed this simulation to estimate the expected variation of Δ*t*_*arrival*_ and Δ*t*_*deviation*_. Virtual artifact arrival times are randomly generated with microsecond resolution. To simulate the impact of the finite resolution of the fiducial point detection, each virtual timestamp was rounded up to the nearest 2 ms in the ECG signal and up to the nearest 16 ms in the PPG signal. The contributions of the two sawtooth artifacts (*Delay*_*sync*_) were also simulated using the amplitude and periods derived from the Kalman filters. Subjectivity involved with measurements made during the hardware profiling experiment was incorporated into the model by assuming that the true artifact location may have been ±1 sample away from the recorded location for Δ*t*_*arrival*_ and up to ±4 samples away for Δ*t*_*deviation*_.

This simulation results in probability density functions that represent the relative distribution of possible latent PAT values as a function of the observed PAT measurement. After validating these transfer functions against experimental observations from our hardware profiling experiment, we use maximum likelihood estimation (MLE) to approximate the effect as a function of the distribution of Δ*t*_*arrival*_ and Δ*t*_*deviation*_. This allows us to estimate *PAT*_*physiological*_ based on a window of preceding *PAT*_*corrected*_ values.

### 3.8 Calculation of Final PAT Distributions

While the methodology described up until this point can be used to correct for system-related timing errors in PAT measurements, we applied a few modifications to calculate robust PAT distributions for different age groups. We first synchronized the ECG and PPG signals by correcting for delays introduced to the PPG timestamps. After subtracting *Delay*_*bu f f er*_ and *Delay*_*filter*_ from each PPG timestamp, we corrected for synchronization error by subtracting half of the amplitude of each sawtooth artifact rather than the exact error estimated by the Kalman filter. Doing so reduced the impact of the filter’s variable performance across the entire dataset. Once the ECG and PPG signals were synchronized, we used the naïve PAT measurement approach described in Section 2.1. In other words, we were able to measure PATs by assessing the time between each R-peak and the following beat onset in the PPG signal. This approach of inter-signal synchronization followed by naïve PAT measurement had the advantage that PAT measurements could be generated for nearly every heartbeat in our database.

To meaningfully compare the PAT measurements before and after correction, we collated values from all patients within the same age groups. We adopted the age bins defined by Eytan et al.^54^, but we split some of the groups below 1 year to make their sample sizes more comparable to those of older age groups. Note that patients with long admissions potentially contributed to multiple age bins. Once the PAT measurements were grouped by age, we inspected the distributions and noticed that a small number of implausibly large values were present due to random failures of the algorithms used to locate fiducial points. The upper end of each distribution was trimmed to remove these physiologically implausible values.

## 4 Results

### 4.1 Dataset Size

The entire dataset comprised 1.42 million hours of overlapping ECG and PPG signals collected from 10,488 unique patients. These signals contained over 7.54 billion heartbeats. When we first began processing our dataset, we quickly noticed a bimodal distribution in *PAT*_*beatmatched*_. Upon further investigation, we discovered that the modes could largely be attributed to the change in data collection hardware that was made in 2022 at the study hospital. The peak of the distribution of *PAT*_*beatmatched*_ was 1,567 ms prior to the hardware change and 1,227 ms thereafter. This indicated that the properties of the system-related timing errors changed significantly when the hardware was upgraded.

After this realization, we stratified our analyses according to two epochs: pre- and post-hardware change. We did not know the exact date that hardware was upgraded at each bedspace, so we conservatively excluded all data from 2022 from subsequent analysis to avoid potential cross-contamination. Table 1 reports the quantity of data included in the two epochs. The pre-2022 data epoch consists of 1.10 million hours of data from 7,317 patients, while the post-2022 data epoch consists of 0.32 million hours of data from 2,304 patients. These recordings equate to over 5.8 billion and 1.6 billion heartbeats, respectively.

**Table 1.** A comparison of timing system properties across different data epochs of our dataset.

| Dataset | Pre-2022 | Post-2022 |
| --- | --- | --- |
| Monitors | Philips MP70 | Philips MX750 |
| Interface | IntelliBridge X2 | IntelliBridge X3 |
| Unique Patients | 7,317 | 2,304 |
| Hours of Data (hrs) | 1.10 million | 0.32 million |
| Number of Beats | 5.89 billion | 1.65 billion |
| R-Peak Temporal Resolution (ms) | 2 | 2 |
| PPG Onset Temporal Resolution (ms) | 16 | 16 |
| Median $PAT_{beatmatched}$ (ms) | 1,567 | 1,227 |
| Sawtooth 1 Period (s) | 55.2 | 65.5 |
| Sawtooth 1 Amplitude (ms) | 35.9 | 37.4 |
| Sawtooth 1 Slope (ms/min) | 30.7 | 39.3 |
| Sawtooth 2 Period (s) | 183 | N/A |
| Sawtooth 2 Amplitude (ms) | 12.4 | N/A |
| Sawtooth 2 Slope (ms/min) | 4.1 | N/A |
| $Delay_{buffer} + Delay_{filter}$ (ms) | 1,301 | 967 |
| $Delay_{buffer}$ (ms) | 580 | ? |
| $Delay_{filter}$ (ms) | 721 | ? |

### 4.2 Characterization of Errors

Table 1 compares the two data epochs according to key characteristics in our PAT measurement pipeline. During these experiments, 69% of the total dataset was filtered due to low signal quality. A small percentage of these windows were discarded due to being unable to detect any peaks in the ECG (1.8%) or PPG (0.02%). Most windows were discarded due to having large gaps in the data (19.4%), a standard deviation for either the ECG or PPG IBIs greater than 300 ms (21.4%), or a difference in mean IBI between the ECG and PPG greater than 200 ms (26.4%).

Nevertheless, the fact that we were still left with billions of heartbeats gives us reasonable confidence in our results. It is also important to note that the effective resolution of PPG fiducial points remained consistent across the two epochs (2 ms for ECG, 16 ms for PPG), showing that this observation was not a peculiarity with a single hardware system. We elaborate on the rest of the timing errors and delays below.

#### 4.2.1 Clock Drift Artifacts

As we investigated the discrepancy between the two data epochs, we identified that they contained a different number of superimposed sawtooth artifacts; the pre-2022 data epoch contained two superimposed artifacts, while the post-2022 data epoch only contained one. Since we used a Kalman filter to characterize the slope and period of each sawtooth cycle independently, we examined the consistency of their shape as a way of confirming that this was a hardware-related phenomenon.

Figure 6 shows the distributions of the sawtooth cycle characteristics for the various artifacts in our dataset. The typical cycle in the dominant sawtooth artifact during the pre-2022 data epoch had a period of 55.2 ± 11.0 ms and an amplitude of 33 ± 9.0 ms (Figure 6, Panel A). The corresponding measurements during the post-2022 data epoch were 59.4 ± 14.9 ms and 33.7 ± 13.5 ms respectively (Figure 6, Panel B). Note that we have reported each distribution’s mode and standard deviation with the expectation that the sawtooth characteristics should be consistent within a given epoch. Since we did not impose these constraints for consistency with our Kalman filter, individual cycles may have varied in slope due to physiological phenomena and noise. Still, the tightness of the observed period and amplitude distributions strengthened our confidence that the sawtooth clock drift artifacts were present throughout the data collection period.

**Figure 6.**
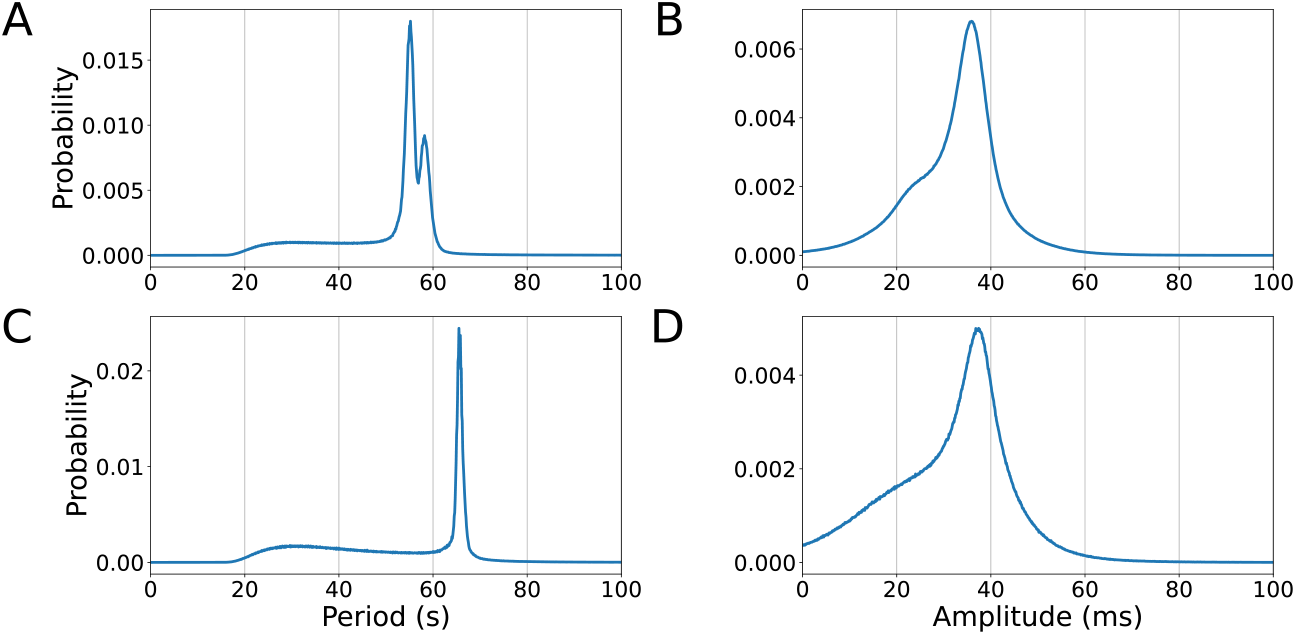
The distributions of various sawtooth cycle characteristics estimated by the Kalman filter: (A) periods across the pre-2022 data epoch, (B) amplitudes across the pre-2022, (C) periods across the post-2022 data epoch, and (D) amplitudes across the post-2022 data epoch.

Once the first sawtooth artifact was removed from the pre-2022 data epoch, it became apparent that there was a secondary sawtooth artifact. The amplitude of this second sawtooth was small compared to the measurement noise, so the Kalman filter was not as effective at measuring its properties. Visual assessment of the sawtooth revealed a period of approximately 180 s and an amplitude of approximately 12 ms (Figure 2, Panel D). We refined these values using a combination of manual fine-tuning and iteration, so we cannot guarantee that these properties are representative of the secondary sawtooth present throughout the pre-2022 data epoch. We report the properties of the sawtooth artifacts in Table 1 and use them in our subsequent estimation of *Delay*_*sync*_.

#### 4.2.2 Hardware Profiling

##### Pre-2022 Data Epoch

Due to logistical limitations, we were only able to profile the hardware during the pre-2022 data epoch. During this period, signals were collected using Philips IntelliVue MP70 patient monitors and X2 IntelliBridge device interfaces. We introduced and observed a total of 136 individual artifacts in our hardware profiling experiment. Observation of the experimental results was more challenging when points of interest coincided with certain parts of the waveform morphology, so only high-confidence measurements were recorded. Δ*t*_*deviation*_ also had a higher variance than Δ*t*_*arrival*_ since the moment of arrival of the artifact in the PPG signal was much easier to locate precisely than the moment when the beat morphology began to deviate from the repeated template of the synthetic signal. Therefore, the results of our hardware profiling experiment include 113 observations (83.0%) of Δ*t*_*arrival*_ and 85 observations (62.5%) of Δ*t*_*deviation*_.

Figure 7 shows the results of our profiling experiment. Observations of Δ*t*_*arrival*_ had a mean and standard deviation of 1,324.2 ± 11.2 ms (Figure 7, Panel A), while observations of Δ*t*_*deviation*_ had a mean and standard deviation of 603.7 ± 29.7 ms (Figure 7, Panel D). The corresponding simulated distributions are shown in Panels B and E respectively, with the simulated distributions having similar variance to the observed data. However, note that the latent values of these measurements are not located at the mean of the observed and simulated distributions due to the presence of *Delay*_*sync*_ and the asymmetrical nature of the measurement errors. Visualizations of the fit between the theoretical and the measured distributions are shown in Panels C and F according to MLE. Our conclusion from this experiment was that the time offset in the PPG due to *Delay*_*bu f f er*_ +*Delay*_*filter*_ was 1301 ms, with 580 ms being attributed to *Delay*_*bu f f er*_ and the remaining 721 ms being attributed to *Delay*_*filter*_.

**Figure 7.**
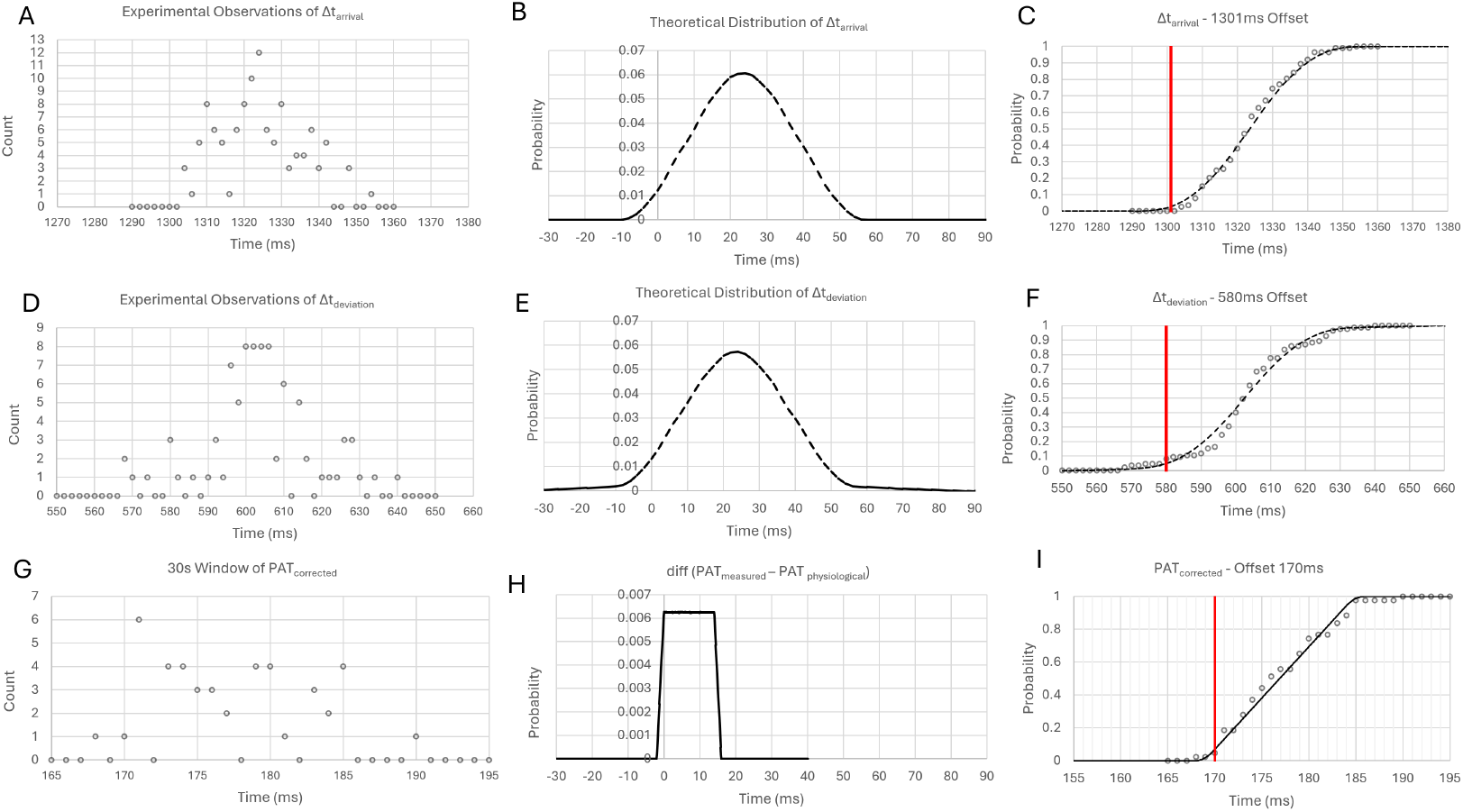
(A) The distributions of observed Δ*t*_*arrival*_ from the hardware profiling experiment. (B) The theoretical probability density function of Δ*t*_*arrival*_ generated by the transfer functions between the measurement space and the latent space. (C) The results of MLE matching the theoretical distribution with the observed measurements. (D, E, F) The same figures but for Δ*t*_*deviation*_. (G, H, I) The same figures but for *PAT*_*corrected*_ over a 30-second window.

##### Post-2022 Data Epoch

Although we were only able to profile the hardware before the change in 2022, we compared the distribution of PAT values at this point of the pipeline (*PAT*_*beatmatched*_ *−Delay*_*sync*_) across the two epochs to estimate the system-related lag (*Delay*_*bu f f er*_ +*Delay*_*filter*_) in the latter one. The system-related lag for post-2022 data epoch was found to be 967 ms. In the absence of performing further hardware profiling experiments, we were unable to further de-convolve this lag into its individual components.

### 4.3 PAT Distribution Comparison Across Ages

Although we could have reconciled the discrepancies between the two data epochs under the assumption that the underlying distribution of *PAT*_*physiological*_ remained unchanged, we opted against this option to prioritize precision. Therefore, we examined the impact of our methodology on PAT measurements drawn strictly from the pre-2022 data epoch. This dataset consists of 5.89 billion heartbeats from 7,317 patients. By characterizing the timing errors and then synchronizing the entire dataset at once so that we could measure PAT naïvely, we did not have to be as selective about filtering our data. After discarding the physiologically implausible PAT values, we were still left with 5.53 billion heartbeats.

Figure 8 illustrates the distributions of *PAT*_*physiological*_ across different age groups. Across all age groups, we observed that the naïve PAT measurements followed a fairly uniform distribution from 0 ms up to 350–400 ms, after which there was a steep drop-off. For the age groups above 4 years, there is a notable peak in the distributions near 700 ms. We believe that these measurements are a consequence of complex interactions between slower heart rates and beat alignment. Past works have discarded such values as outliers^25,36^, resulting in distributions with physiologically plausible averages. Still, the uniformity of these distributions illustrates the remaining flaws of the naïve measurement approach.

**Figure 8.**
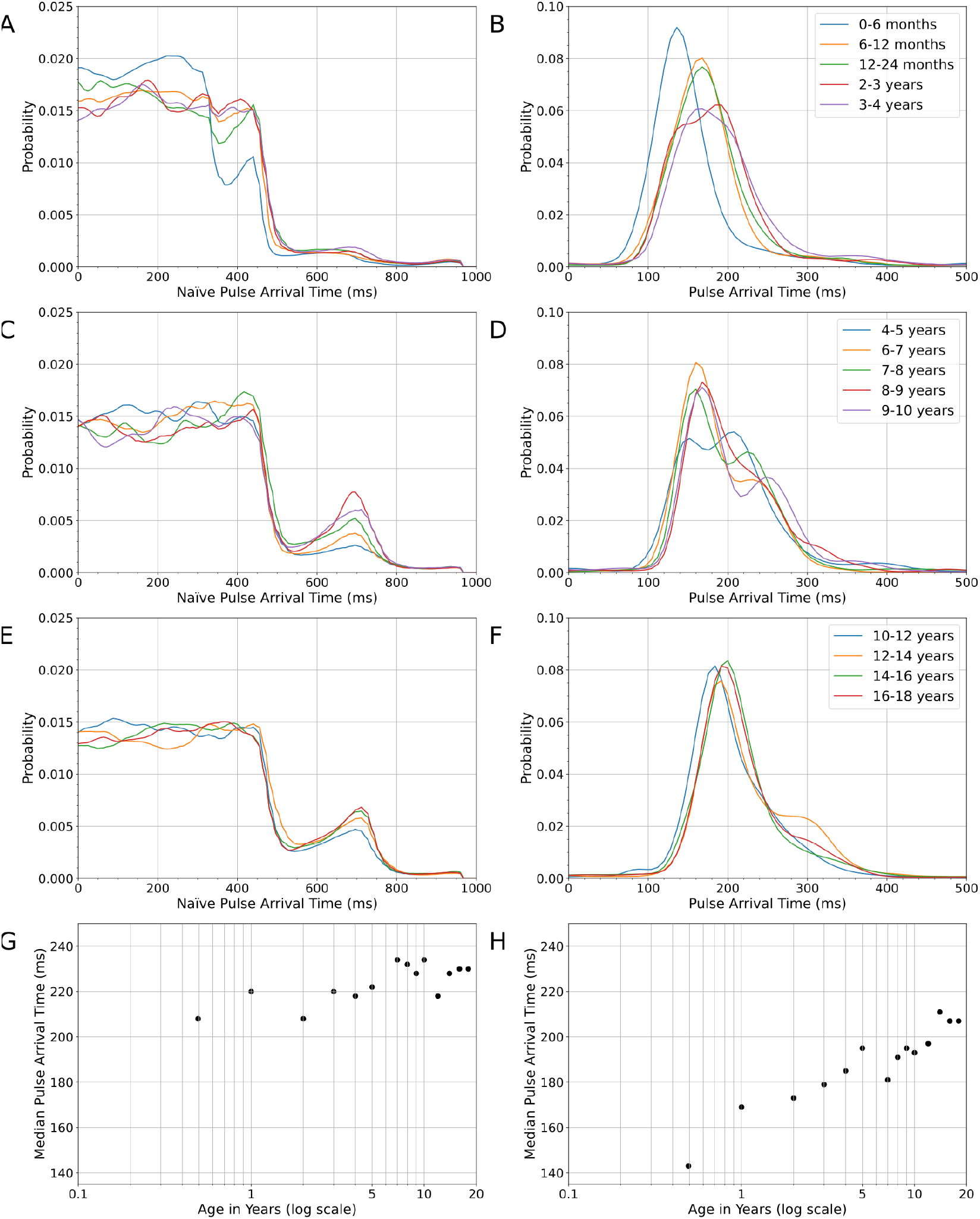
Various comparisons of PAT measurements (left) before and (right) after applying our methodology to correct for system-related timing errors. The first three rows compare the probability distribution functions of different age groups, while the bottom row compares the median PAT values across a logarithmic scale.

After applying our methodology to remove system-related timing errors, we observed more logical normal distributions. Some of the distributions for the intermediate age groups exhibited two modes. We speculate potential explanations for these modes in Section 5.4, but the fact that they are closer together than those observed in the naïve measurement distributions gives us confidence that they are not due to measurement error. The logarithmic increase in median PAT across age groups further reinforces our confidence in these measurements. As patients become older, the separation between ECG and PPG measurement sites becomes greater due to changes in height and limb length. Since it takes more time for the pulse to travel greater distances, these anatomical parameters play a major role in PAT measurement.

## 5 Discussion

### 5.1 Summary of Key Findings

Our work highlights a multitude of confounders that can complicate PAT measurement using standard equipment in the ICU. ECG and PPG signals are often sampled using two separate devices, each of which typically operates with its own clocks before being collated by downstream monitoring hardware. Since the hardware is designed primarily for real-time signal visualization, systems often include proprietary mechanisms such as digital filtering and buffering to pre-process the waveforms. These stages can introduce a time offset between ECG and PPG that is large enough to become a significant confounder for PAT measurement.

Meanwhile, prior work has previously reported sawtooth artifacts in PAT timeseries data but without a consistent explanation^42–44^. We posit that these artifacts are the result of relative clock drift and periodic resynchronization between signals, producing timing errors that accumulate until they are periodically re-synchronized. In fact, we observed that multiple sawtooth artifacts can be superimposed on one another when resynchronization happens in multiple stages, as was the case in our pre-2022 data epoch. Finally, many studies have examined how quantization leads to differences in HRV metrics calculated with ECG and PPG with different sampling rates^61–64^. To the best of our knowledge, we are the first to explicitly model and correct measurement errors resulting from the finite temporal resolution of fiducial points across two signals in the context of PAT measurements.

Medical device manufacturers would ideally disclose any timing delays and details of filters applied to ECG and PPG signals, as that would make it easier for researchers to address the challenges identified in our paper. While this may happen in the future, the reality now is that this information is largely undocumented or considered to be proprietary. Therefore, our work presents a procedure that researchers can apply in order to precisely measure PAT despite the presence of hardware-related timing errors. The properties of the timing errors we identified in our dataset are likely to vary across hardware systems. Nevertheless, the magnitude of these errors relative to the PAT measurements we report in Section 4.3 emphasizes the importance of acknowledging and addressing these confounders.

### 5.2 Implications for Real-Time BP Estimation

ICU patients may exhibit rapid fluctuations in BP that require immediate detection and intervention. This is typically accomplished using an intra-arterial catheter that imposes risk of injury if improperly placed or jostled^65^. Many children who are at risk of deterioration but who are not acutely unstable do not have such catheters, even in an ICU setting, and the utilization of an intra-arterial catheter is only plausible in high-acuity settings such as the OR or ICU. Since hemodynamic fluctuations can occur in these situations where invasive intra-arterial monitoring is not being used, a measure such as PAT could act as an early warning system to signal the clinician to measure blood pressure more frequently or more accurately by providing noninvasive insights into PWV and BP^5–8^. Calculating PAT using signals that can be collected using noninvasive sensors may yield a safer and more broadly available alternative to invasive monitoring, especially since ECG and PPG are routinely and continuously monitored in various clinical settings.

Achieving real-time PAT measurements requires making a few adjustments to our proposed approach so that it can be applied prospectively rather than retrospectively. Quantifying and correcting system-related lags can be done using a one-time hardware profiling experiment, but correcting the time-varying errors introduced by the sawtooth artifacts requires continuous analysis. If the amplitude and period of each sawtooth can be precisely estimated with a one-time retrospective analysis, only the phase must be inferred in real time. While our Kalman filter algorithm treated each sawtooth cycle independently so that we could examine the consistency of the phenomena across our dataset, a real-time implementation could rely on information from previous cycles to estimate the phase at any given moment. Once the sawtooth artifacts were removed, we used MLE to account for measurement errors and estimate the latent physiological PAT values. There will be a trade-off between window size and confidence in the resulting output, with larger windows allowing for better fitting at the cost of increased lag in the final output. Longer windows may also be increasingly subject to the underlying physiological variability of the latent PAT signal.

Real-time BP estimation using PAT measurement relies on the correlation between the two variables. Prior work has reported on this relationship with mixed results^5,18,32,66,67^. Due to the correlation between PAT measurements and HR^21,27,68^, we suspect that many studies using PAT for BP estimation may actually be overly reliant on the correlation between HR and BP. Beyond the physiological causes for this correlation, our work revealed that naïve assumptions about signal synchronization resulted in a far stronger correlation between HR and PAT due to the phase interactions related to incorrectly matched beats between the signals. Many researchers have been able to bypass this problem to some extent by training and testing models on individual patients rather than across them or by utilizing patient-specific variables such as baseline HR to improve results^36^. Other groups have explicitly accounted for the correlation between HR and BP in their models^19,22^. While these approaches may be suitable in some situations, creating a truly generalizable BP estimation technique in high-acuity settings will require further consideration.

### 5.3 Implications for Deep Learning Models

The proliferation of multimodal datasets such as MIMIC^14^, WAVES^10^, MOVER^15^, and VitalDB^13^ has led to a surge of research in pursuit of accurate deep learning models to predict clinical outcomes. In addition to BP prediction, several groups have used PAT and other multi-waveform features as inputs to predict sepsis^69^, septic shock^70^, depth of anesthesia^71^, and patient decompensation^72^. Raw ECG and PPG signals have also been used as model inputs to predict blood pressure^73–83^ and generate foundational models for non-invasive physiological sensing^84–87^.

While deep learning models may learn to account for constant and time-varying timing offsets inherent to a single dataset with enough training, they will likely fail to generalize to other hardware systems with different hardware offsets and different synchronization artifacts. Training these models on datasets sourced from multiple sites is equally problematic, as we cannot necessarily expect a deep learning model to reconcile the differences across datasets on its own. We have demonstrated that even data from a single site may contain different data epochs, each with its own characteristic timing errors. Therefore, we argue that quantifying and correcting inter-signal timing errors is necessary for robust model generalization.

Beyond models involving ECG and PPG, the techniques described in this paper could potentially be used to measure and correct the relative timing errors between other physiological signals that contain information about cardiac activity. For instance, the electrical activity of the heart can also introduce subtle artifacts into electroencephalography (EEG)^60^, so the beat-matching algorithm can be applied to identify how this signal lags relative to the PPG or ECG. This approach, combined with the other algorithms we present for correcting clock drift artifacts and quantization errors, can then be used to correct inter-signal timing errors, opening the door to large retrospective analyses that rely on inter-signal synchronicity.

### 5.4 Caveats on Our PAT Distributions

Previously reported ranges in PAT have varied significantly^21,88,89^. The disagreement may be partly attributed to varied definitions for PAT. Some groups measure PAT as the time between the R-peak in the ECG and the pulse peak in the PPG, while others rely on pulse onset in the PPG. PAT and PTT may have also been used interchangeably between different groups, with the latter often describing the time difference between pulse arrival at the finger versus the foot^90^. Regardless, the discrepancy across studies is significant enough to convey that PAT is not being objectively measured between datasets, research groups, and hospitals. The potential for relative timing errors between the ECG and PPG signals has either been overlooked or assumed to be negligible^91^.

While we have attempted to be specific about how PAT is defined and measured in this paper, our comparison of PAT measurements across age groups still suffers from some limitations and open questions:

1. **Patient Health Status:** Because our dataset was drawn from critically ill children in an ICU, our ranges are unlikely to be representative of the general population. Children may be admitted to the ICU for a broad range of diagnoses and underlying medical conditions that can alter normal physiologic states and consequently influence the PAT (e.g., congenital heart disease). We also did not have information about medical interventions that were administered to the patients to help control their BP, which likely influenced their PAT beyond the physiological variability that would be present in healthy children.
2. **Measurement Site:** Because it takes more time for the pulse to travel further, PAT is inherently correlated with the distance between the two locations where the pulse is detected^92,93^. While ECG leads are typically placed at consistent positions across the chest, the location of the PPG sensor may vary depending on the circumstances. PPG sensors are typically placed on adults’ fingertips, yet it is not always feasible to do the same with children. In some of these cases, the PPG is occasionally placed on another extremity like a toe or ear. Unfortunately, the PPG monitoring site was not recorded in our dataset.
3. **Patient Height:** Even when measurement sites are kept consistent, patients of varying heights are likely to exhibit different PATs because of changes to the arterial path length being characterized. While our analysis accounts for this confounding factor to a degree by using age as an indirect proxy for height, more could be done to disambiguate the influence of limb length and other demographic variables.
4. **Global Correction of Sawtooth Artifacts:** The algorithm we used to globally correct for the sawtooth artifacts when calculating the PAT distributions was simplistic in the sense that it only shifted the mean but did not alter the variance of the data. We performed a Monte Carlo simulation to confirm that the shift in the distributions’ standard deviations was well below 1 ms using the techniques described by Frenkel and Kirkup^94^, but deeper investigation should be conducted to support this belief.
5. **Signal Timestamping:** We considered signal timestamps to be at the end of each sample since that is more logically consistent with the physical and temporal reality of the sampling process, yet we acknowledge that this decision requires further justification. Assuming that the timestamps describe the beginning of each sample would still generate meaningful PAT measurements, but such an assumption would shift every *PAT*_*corrected*_ value by +14 ms in our dataset. It is important to note, however, that the values for *PAT*_*physiological*_ calculated in this work would be identical irrespective of how samples are assumed to be timestamped. This is because the output of the MLE that generates the latent PAT signal would be almost identical.

Despite these limitations, the significant scale of our dataset gives us reasonable confidence that the methodology we have presented yields more clinically meaningful data.

### 5.5 Future Work

Beyond addressing the limitations that have already been mentioned with respect to our calculated PAT distributions, there are many other opportunities for refinement and exploration. For example, our algorithm for tracking sawtooth artifacts treated each cycle independently. While this allowed us to support the consistency of the phenomenon, the algorithm could be improved to address discrepancies across cycles. One way to do this would be using the parameters inferred from previous cycles to serve as the initial guesses for future ones.

Lastly, having access to a large-scale dataset with high-precision PAT measurements lends itself to revisiting the correlation between PAT and BP in critically ill children. We now have access to billions of precisely measured PATs, and about 40% of that data is also covered by simultaneously collected arterial BP waveforms that can be used as a ground truth for BP. This dataset should be sufficient to enable robust statistical analyses and the development of more accurate models for understanding the relationship between PAT and BP.

## 6 Conclusion

Timestamps assigned to PPG signals may be impacted by several factors including clock drift, buffering, and digital filters. Traditional approaches to PAT measurement implicitly assume that PPG signals are precisely synchronized with other physiological waveforms. We demonstrate that these timing errors can be quantified and corrected via experimental profiling of the bedside monitoring equipment and the application of a novel PPG synchronization algorithm. We tested these algorithms on a large retrospective dataset comprising nearly 2 million hours of signals collected from a pediatric ICU. We showed that timing errors were indeed caused by system hardware and software, and then we demonstrated that applying our methodology led to more clinically meaningful PAT measurements. It is our hope that other researchers interested in using similar multimodal datasets acknowledge the importance of the timing issues highlighted by our work.

## Data Availability

All data produced in the present work cannot be made publicly available because they contain personal health information.

## Code availability

Data is collated in an open-source AtriumDB database and analyzed using Python 3.10. The code used to analyze the dataset is available upon reasonable request from the corresponding author.

## Author Contributions

I.R., A.S., A.M., and A.G. conceived the research objectives and experiments. I.R. and A.G. conducted the experiments, while I.R., A.S., W.D., A.M., and A.G. analyzed the results. B.N. conducted the hardware profiling experiments. All authors reviewed the manuscript.

## Footnotes

1 https://github.com/neuropsychology/Neurokit

2 https://github.com/PIA-Group/BioSPPy

